# A Model-Based Strategy on COVID-19 Vaccine Roll-out in the Philippines

**DOI:** 10.1101/2022.05.27.22275675

**Authors:** Rey Audie S. Escosio, Olive R. Cawiding, Bryan S. Hernandez, Renier G. Mendoza, Victoria May P. Mendoza, Rhudaina Z. Mohammad, Carlene P.C. Pilar-Arceo, Pamela Kim N. Salonga, Fatima Lois E. Suarez, Polly W. Sy, Thomas Herald M. Vergara, Aurelio A. de los Reyes V

## Abstract

Coronavirus disease 2019 (COVID-19) is an infectious disease caused by severe acute respiratory syndrome coronavirus 2. Millions of people have fallen sick, and some have died due to this affliction that has spread across the globe. The current pandemic has disrupted normal day-to-day human life, causing a profound social and economic burden. Vaccination is an important control measure that could significantly reduce the incidence of cases and mortality if properly and efficiently distributed. In this work, an age-structured model of COVID-19 transmission, incorporating an unreported infectious compartment, is developed. Three age groups are considered, namely: *young* (0-19 years), *adult* (20-64 years), and *elderly* (65+ years). The transmission and reporting rates are determined for each group by utilizing the number of COVID-19 cases in the National Capital Region in the Philippines. Optimal control theory is employed to identify the best vaccine allocation to different age groups. Further, three different vaccination periods are considered to reflect phases of vaccination priority groups: the first, second, and third account for the inoculation of the elderly, adult and elderly, and all three age groups, respectively. This study could guide in making informed decisions in mitigating a population-structured disease transmission under limited resources.

## 1 Introduction

Coronavirus disease 2019 (COVID-19) is an infectious disease due to severe acute respiratory syndrome coronavirus 2 (SARS-CoV-2). Two years have passed since the World Health Organization characterized the COVID-19 outbreak as a pandemic [11], with over 468 million confirmed cases and just over 6 million reported deaths globally at the time of writing [10]. Its emergence has disrupted not only public health but also the global economy. Unfortunately, with the highly contagious Omicron variant driving the current increase of reported cases around the world [10], this pandemic is still not over.

To analyze the transmission dynamics of this infectious disease, mathematical modeling has extensively been used and has played an essential role in predicting, assessing, and controlling outbreaks [44]. Numerous compartmental epidemiological models, such as SIR (susceptible-infectious-recovered), SIRS (susceptible-infectious-recovered-susceptible), and SEIR (susceptible-exposed-infectious-recovered) models, have been developed in the past decades, to represent the transmission of infectious diseases [16, 17, 25, 32]. Moreover, to properly identify interventions that can prevent outbreaks, one needs to take into account the social structure and mixing patterns, which vary across countries in different stages of development and with different demographics, in the epidemic model [39]. In the Philippines, early studies on COVID-19 revealed that while only eight percent of the population are in the age group 60 and over, they account for the majority of all COVID-19 deaths so far; and while individuals below age 60 dominate the reported cases, they account for only a third of total deaths [2, 12, 43].

Vaccine is a biological preparation that stimulates one’s immune response against a particular disease. Vaccine administration known as vaccination is an effective method designed to prevent an infectious disease [5]. To reduce the risk of contracting COVID-19, several vaccines have been approved worldwide for public use [1]. However, due to global demand and limited supply of vaccines against the virus, analysis of priority in the vaccine roll-out is vital.

Several studies use optimal control theory to investigate compartmental models in epidemiology. In particular, Gaff and Schaefer (2009) focused on finding the optimal response balancing vaccination and treatment strategies that will minimize the number of infected individuals with variations of standard SIR, SIRS, and SEIR epidemiological models [24]. In addition, Hansen and Day (2011) extended their work on the optimal control of a basic SIR epidemic model and obtained policies for an isolation-only model, a vaccination-only model, and a combined isolation–vaccination model [26]. For the COVID-19-specific study, Kantner and Koprucki (2020) provided a study for the epidemics with purely non-pharmaceutical interventions based on an extended SEIR model and optimal control theory [29]. Moreover, Djidjou-Demasse et al. (2020) investigated optimal COVID-19 epidemic control via social distancing and lockdown measures until the deployment of vaccines [20]. Additionally, Obsu and Balcha (2020) used optimal control to minimize the number of exposed and infected populations that took into account the cost of implementation [36]. In the Philippine setting, Estadilla et al. (2021) considered the impact of vaccine supplies and delays on the optimal control of the COVID-19 pandemic [23]. Furthermore, Olivares and Staffetti (2021) considered optimal control to suggest a suitable schedule for vaccination and testing policies to control the spread of COVID-19 [37]. Finally, age-structured non-pharmaceutical interventions for optimal control of the COVID-19 epidemic were established by Richard et al. in the Burkina-Faso, France, and Vietnam settings [42].

In this work, we focus on the National Capital Region (NCR), which is the most densely populated region and the center of the economy of the Philippines [8]. Our study generally aims to develop an age-structured model that could describe the spread of COVID-19 in the NCR and identify effective control strategies based on optimal control theory. Specifically, this work aims to consider a dynamic model incorporating the effects of age-dependent susceptibility to COVID-19 infection in the NCR; obtain time-dependent optimal vaccination policies based on priority groups in different periods of vaccine roll-out; and assess the corresponding allocation of vaccines to each group. To the best of our knowledge, this is the first study that applies optimal control theory to a COVID-19 age-structured model in the Philippines. Since testing and contact tracing of suspected COVID-19 cases in the Philippines were lagging [27], our model incorporates possible unreporting in the different age groups. The capacity of daily vaccine administration is also considered to identify the prioritization of vaccination in different age groups so that the number of infections is minimized.

## 2 Methodology

### 2.1 Age-Structured Model

In this work, a modified deterministic SEIR compartmental framework is used to model the progression of COVID-19 in the National Capital Region (NCR), Philippines. We consider the total population (*N*) as a sum of the four epidemiological classes: susceptibles (*S*), exposed (*E*), infected (*I*), and recovered (*R*). Individuals from the susceptible population can be exposed to the disease from interaction with infected individuals at a transmission rate *λ*. Upon acquiring infection, they move to the exposed class and stay in this stage for an average period of, 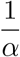 where *α* is the latency rate. In our model, we assume that the *incubation period*, which is the time period from acquiring the virus until developing symptoms, has the same duration as the *latent period*, the period from acquiring the virus until becoming infectious [13, 15, 31, 33, 40]. After this period, the infection worsens, the exposed individual develops infectivity, and is then moved to the infectious class.

In this model, the infectious class is subdivided into two. First, the reported subclass *I*^r^ is composed of individuals who are tested and recorded as virus-positive with reporting rate *ρ*. The second is the unreported subclass *I*^u^, which considers infectious individuals who are not reported due to the absence of testing brought about by socio-economic burden, under-reporting, and/or misreporting. Taking into account the possible transmission contributions of these two subclasses, we consider the force of infec-tion 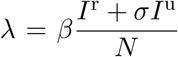, where *β* is the transmission rate and *σ* is a multiplicative factor denoting relative infectiousness of *I*^u^. Note that *I*^u^ includes both asymptomatic and symptomatic infectious individuals who are not in quarantine or isolation. After being infectious, an individual may either progress to the recovered class with a recovery rate *γ*, or die due to the disease with a disease-induced death rate *µ*. The model limits the progression of the disease to a single infection, i.e., recovered individuals are no longer infectious and are immune to reinfection.

It has been observed that COVID-19 cases and risk severity increases with age [19,21]. Since population structure plays a significant role on the disease transmission dynamics [14], each epidemiological class is further divided into three age groups: *young* (0-19 years), *adult* (20-64 years), and *elderly* (65+ years). The grouping is consequential to the following protocols implemented by the Philippine government during the community quarantine placed into effect since March 2020: (1) the group of individuals below 20 years old is comprised mainly of students whose social mixing is impacted by the suspension of face-to-face classes, (2) the majority of the individuals aged 20-64 years old are considered as the working population who are authorized to attend to the basic needs of their families, and (3) the persons who are 65 years old and older are prohibited to go out of their residences, except for unavoidable circumstances [6].

A contact matrix *C* = (*C*_*ij*_) denoting the interactions between an age group *i* with other age groups *j* is incorporated to account for the effect of mixing patterns in the disease transmission [39–41]. Using this, we modify the force of infection *λ*_*i*_ for a susceptible individual in age group *i* as follows

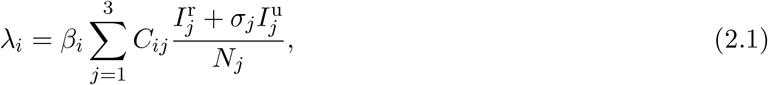

where *β*_*i*_ is the transmission rate of age group *i*, the notations 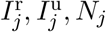 are the reported infectious, unreported infectious, and total population of age group *j*, respectively, *σ*_*j*_ is a multiplicative factor relative to the transmission contribution of 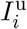 in group *j*, and *i, j* = 1, 2, 3, correspond to each age group. Figure 1 schematically depicts the disease progression for an age group *i*.

**Figure 1:**
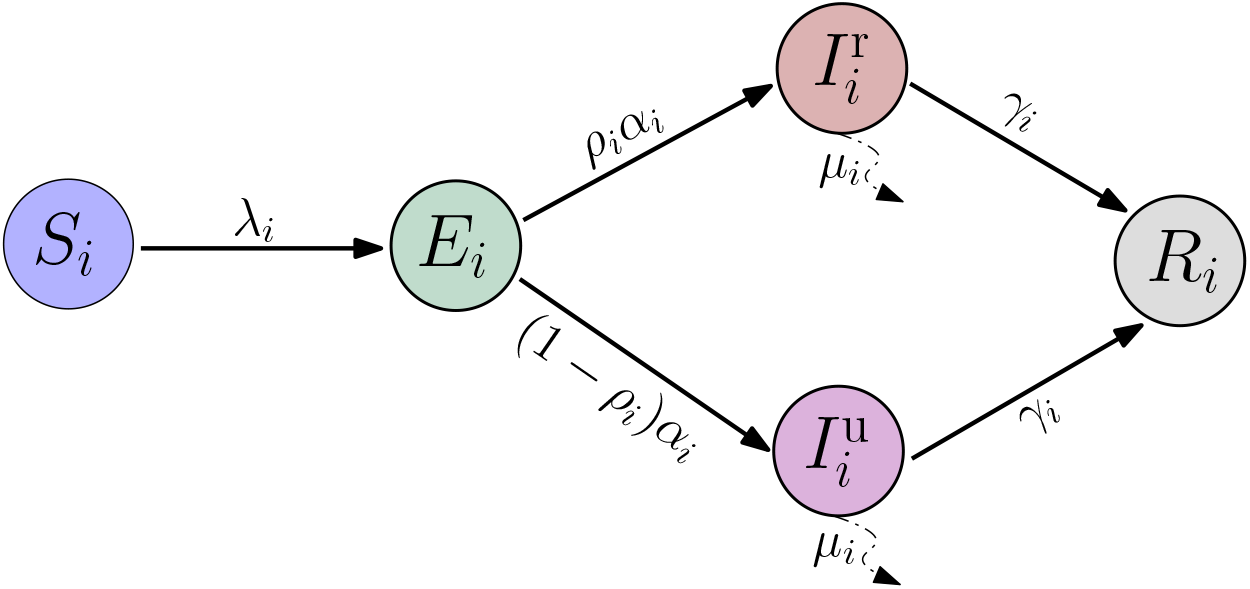
Schematic diagram of the COVID-19 disease transmission model. Compartments in the model are classified into distinct epidemiological classes: Susceptible (*S*), Exposed (*E*), Reported Infectious (*I*^r^), Unreported Infectious (*I*^u^), and Recovered *R*. Each compartment is then stratified into three age groups: Young (0-19), Adult (20-64), and Elderly (65+). The subscript *i* denotes a particular age group.

The disease transmission dynamics can be described by the following set of coupled ordinary differential equations

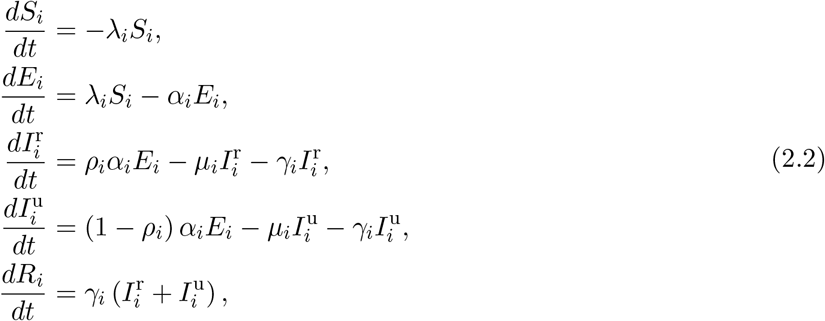

where *λ*_*i*_ is the force of infection, *α*_*i*_ is the latency rate, *ρ*_*i*_ is the reporting rate, *µ*_*i*_ is the death rate, and *γ*_*i*_ is the recovery rate, for a particular age group *i*. Table 2.1 lists the set of parameters, their descriptions and units, including initial values for the state variables of the three different age groups.

**Table 2.1:** Parameters and initial values for the different age groups.

| Parameter | Unit | Age Groups |  |  | Source |
| --- | --- | --- | --- | --- | --- |
|  |  | Young (0-19) | Adult (20-64) | Elderly (65+) |  |
| transmission rate ($\beta$) | 1/day | 0.0056 | 0.0115 | 0.0284 | estimated |
| factor of unreported transmission ($\sigma$) | dimensionless | 2 | 2 | 2 | assumed |
| latency rate ($\alpha$) | 1/day | 0.20 | 0.20 | 0.20 | assumed |
| reporting rate ($\rho$) | dimensionless | 0.2240 | 0.7011 | 0.9980 | estimated |
| death rate ($\mu$) | 1/day | 0.0476 | 0.050 | 0.0476 | [3] |
| recovery rate ($\gamma$) | 1/day | 0.0282 | 0.0305 | 0.0333 | [3] |
| initial susceptible population ($S_0$) | people | 5,073,679 | 8,147,853 | 539,190 | computed |
| initial exposed population ($E_0$) | people | 1,845 | 2,957 | 196 | computed |
| initial reported infectious population ($I_0^r$) | people | 30 | 328 | 42 | [3] |
| initial unreported infectious population ($I_0^u$) | people | 286 | 177 | 0 | computed |
| initial recovered population ($R_0$) | people | 78,193 | 108,425 | 9,237 | [3] |
| initial deceased population ($D_0$) | people | 1,511 | 2,095 | 179 | [3] |
| total population ($N_0$) | people | 5,155,544 | 8,261,835 | 548,844 | [8] |

### 2.2 Data Acquisition

Age-specific case numbers compiled and reported by the Department of Health (DOH) in the Philippines are used in this study. Information from January 1, 2021 until February 28, 2021, on the incidence cases for young, adult, and elderly groups are extracted from the DOH Data Drop [3].

The study considers only the National Capital Region (NCR) as the area of interest due to its soaring number of cases, its high mobility and population density, and most importantly, its significant share of the vaccine allocation. The daily incidence cases for the three different age groups considered are shown in Figure 2(A-C). The biggest share of reported cases is among the adult group with 82.80%, followed by the young group with 9.27%, while the elderly group had the least contribution of only 7.93% of the total cumulative cases since January 1, 2021, as depicted in Figure 2(D). An increasing trend of cumulative cases can also be observed across the three age groups. Note that the adult group constitutes the biggest population share of about 59.16%, and the relative population shares of young and elderly groups are, 36.91% and 3.93%, respectively. However, in terms of infection rate, the elderly group has the highest susceptibility to infection with 3.4%, followed by the adult and young groups, with 2.3% and 0.3%, respectively (refer to Figure 2(E)).

**Figure 2:**
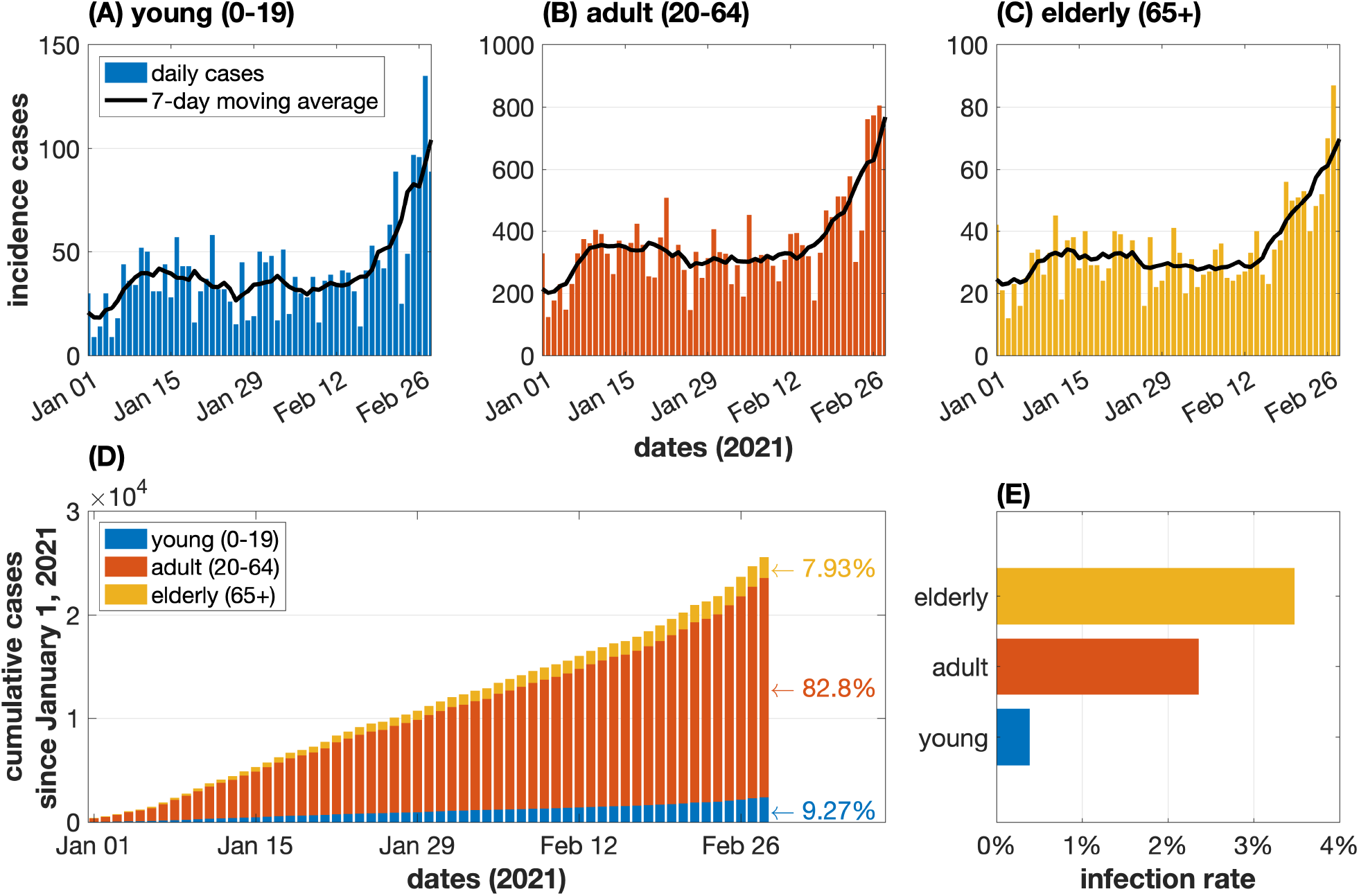
(A-C) Daily reported cases for young (blue), adult (red), and elderly (yellow) group with the corresponding 7-day moving average (black curve). (D) Cumulative cases for the three different age groups from January 1 to February 28, 2021. Adult group comprises the bulk of COVID-19 cases. (E) Infection rate for elderly, adult, and young populations based on cumulative cases until February 28, 2021. The elderly group shows the highest susceptibility to infection.

### 2.3 Contact Matrices

Transmission of an infectious disease such as COVID-19 is influenced by the social structure involving person-to-person interactions which vary by age and locations – at home, at the workplace, at school, or in the community [33, 41]. Our model considers the effect and contribution of the various social mixing in these different locations.

Matrices for household (*D*^*h*^), workplace (*D*^*w*^), school (*D*^*s*^), and community (*D*^*c*^) contact can be obtained from [41] and are shown in Figure 3. The data consists of four 16 *×* 16 matrices binned in 15 five-year age levels (0 *−* 4, 5 *−* 9, …, 70 *−* 74) and a single age group with ages 75 and above. The total of the four matrices when added can represent the whole contact matrix *D* = (*D*_*kl*_), i.e.,

**Figure 3:**
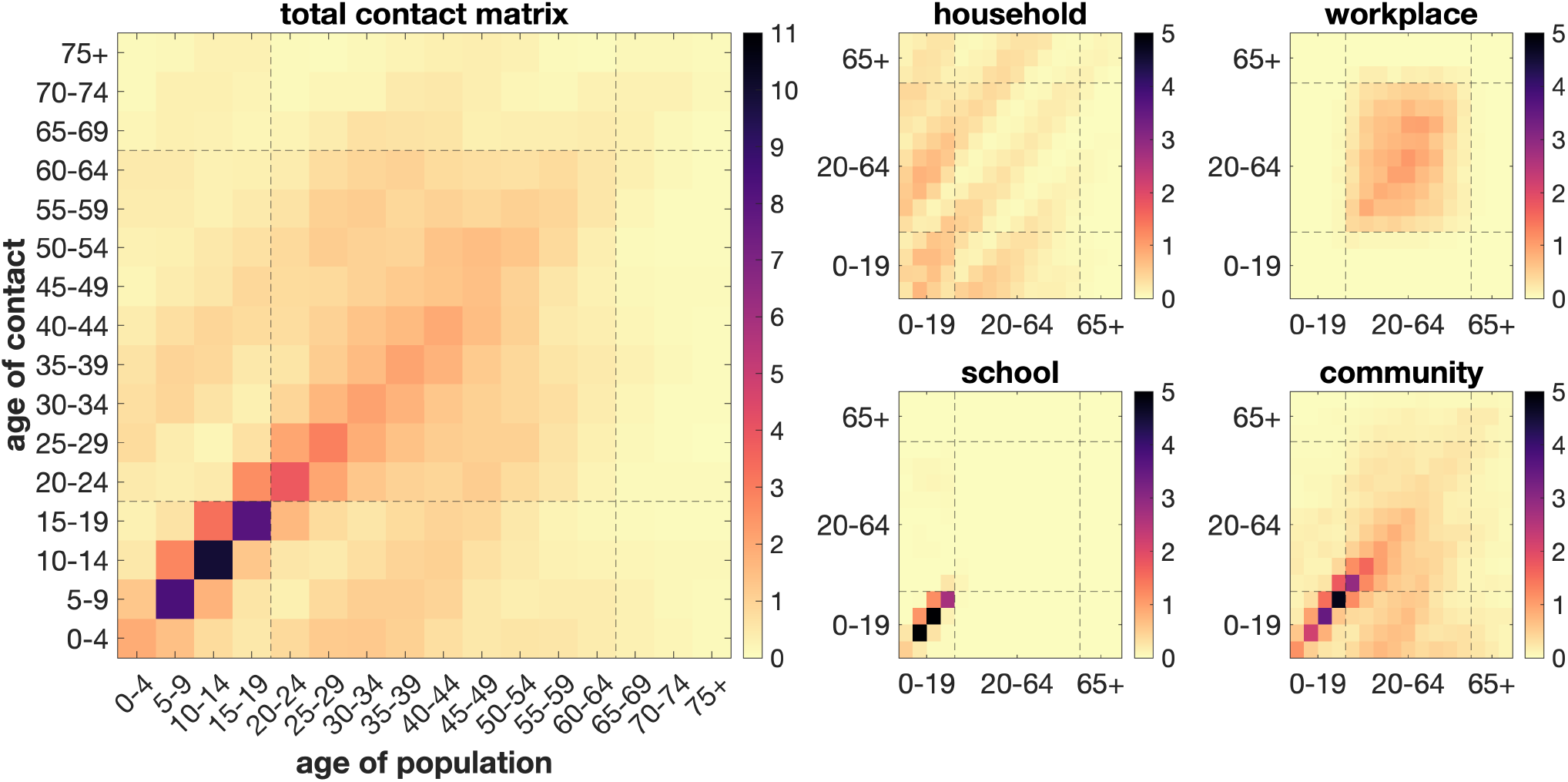
Baseline contact matrices in the Philippines during pre-COVID-19 pandemic showing strong interactions within the same age group in different locations. Figure is reproduced from [41]. For visualization, the shown matrices *D*^*†*^ are flipped upside down, i.e., *D*^*†*^ = 𝕁*D* where 𝕁 is the backward identity.

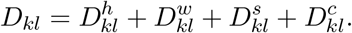

The matrix entries *D*_*kl*_ show a pre-pandemic view of the number of contacts in the *k*th age group of a certain *j*th age group population in the Philippines.

We reduce the size of these matrices from 16 *×* 16 to a mere 3 *×* 3 corresponding to our clumped age groups which are denoted by *C* = (*C*_*ij*_). The process is detailed in Appendix A. Additionally, this 3 *×* 3 total contact matrix *C* = (*C*_*ij*_) can be rewritten accordingly based on [30] to integrate the non-pharmaceutical interventions during the pandemic, i.e.,

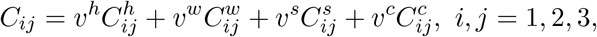

where the coefficients *v*^*h*^, *v*^*w*^, *v*^*s*^, and *v*^*c*^ correspond to the implemented contact interventions in households, workplaces, schools, and communities, respectively. Note that these equate to one if no intervention is implemented, and equate to zero if interactions in such location are entirely halted. One obvious value is *v*^*s*^ = 0 for schools have been closed since the start of the community quarantine. We assign *v*^*h*^ = 1 since household members are maintained to be close contacts. On the other hand, the regulated workforce and community interactions can be assumed to be at 50% or *v*^*w*^ = *v*^*c*^ = 0.50. Applying these coefficients, we get the contact matrices in Figure 4.

**Figure 4:**
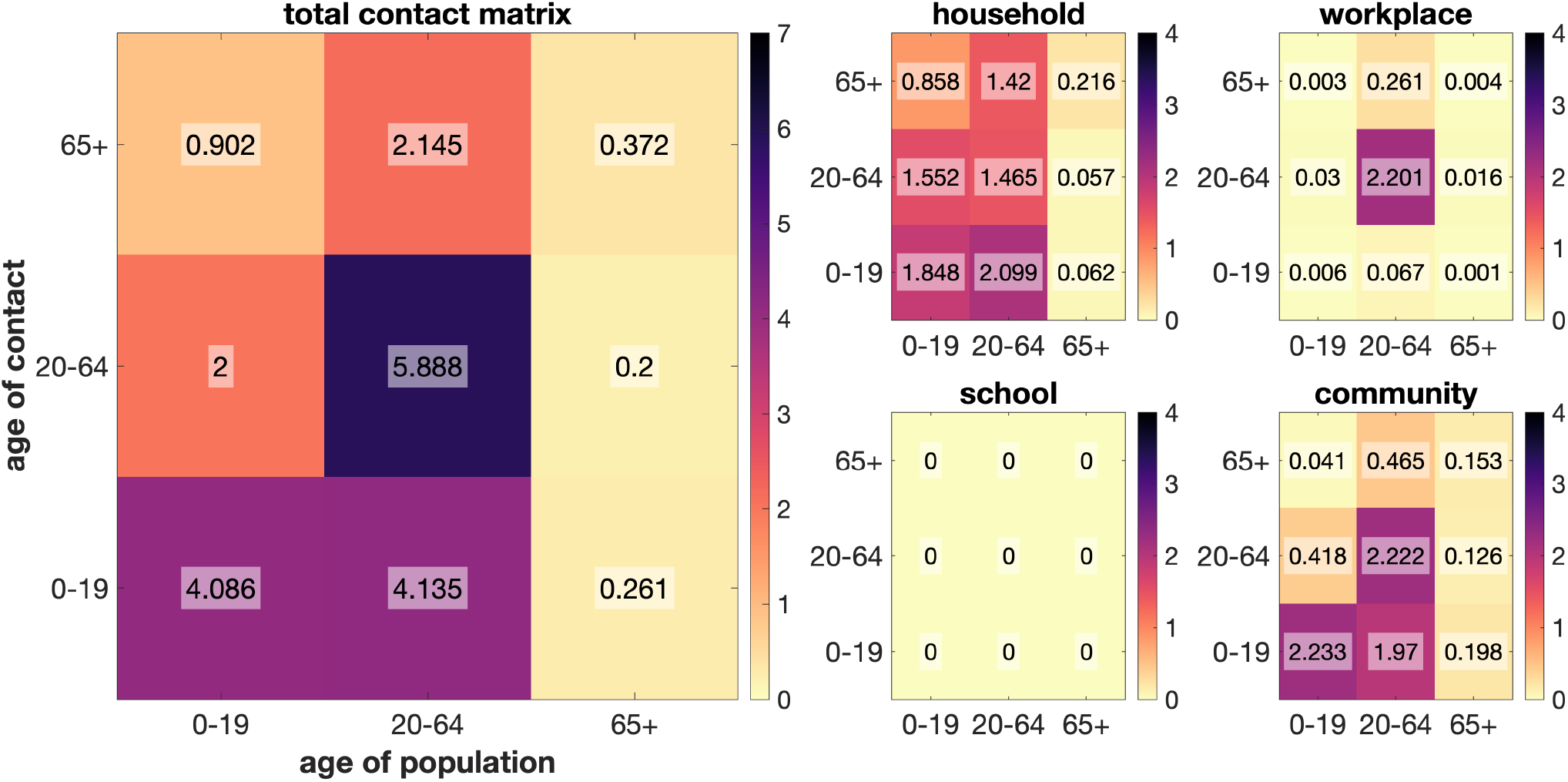
Assumed mitigated 3 *×* 3 contact matrices in the Philippines considering the young, adult, and elderly population groups in total, household, workplace, school, and community context. For visualization, the shown matrices *C*^*†*^ are flipped upside down, i.e., *C*^*†*^ = 𝕁*C* where 𝕁 is the backward identity.

The final contact matrix *C* = (*C*_*ij*_) is shown in the left of Figure 4. For the young compartment, the first row of the contact matrix is [*C*_1*J*_] = [4.086, 4.135, 0.261] where *J* = 1, 2, 3. Even with the interventions brought about by government policies such as the closure of schools, this row contains high values due to continuous contact among the juvenile population in households and communities. As for adults, we have the second row [*C*_2*J*_] = [2.000, 5.888, 0.200] where *J* = 1, 2, 3. For this age group, close contacts are largely observed in the workplace and communities, even though only 50% of the population is allowed to interact. On the other hand, the elderly population has the least contact with all age groups, including itself, as seen in row [*C*_3*J*_] = [0.902, 2.145, 0.372] where *J* = 1, 2, 3. This can be attributed to prohibitions of elderly citizens from activities beyond their homes.

### 2.4 Parameter Estimation

We used the population projection of DOH for 2021 as the initial population for the system (i.e., *N*_0_ = 13, 966, 223). This can be divided accordingly into the young (*N*_1,0_ = 5, 155, 544), adult (*N*_2,0_ = 8, 261, 835), and elderly (*N*_3,0_ = 548, 844) compartments using the available population fractions data. The remaining initial values for the compartments are seen in Table 2.1.

From Table 2.1, the total *E*_0_ is approximated from the result of the 2020 COVID-19 model [22]. On the other hand, the initial reported infectious 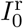, deceased *D*_0_, and recovered *R*_0_ populations are derived from the tallied data in [3] and divided according to the size of the age groups. The initial unreported compartment is approximated as 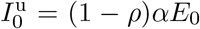, immediately after parameter estimation. As for the initial susceptible population *S*_0_, it can be calculated as 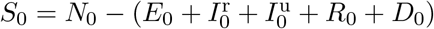.

The multiplicative factor *σ* for unreported infections is held at 2 for all age groups. This presumes that undetected infectious interactions are twice those of the detected. The latency rate *α* is assumed to be 1*/*5, corresponding to 5 days of disease progression from infection to the infectious state. The death rate *µ* and recovery rate *γ* are calculated from the data in [3] since dates of confirmation, death, and recovery are available. Lastly, the transmission rate *β* and reporting rate *ρ* will be identified via parameter estimation. The process of parameter estimation aims to identify selected parameters as a result of minimizing an objective function. In this paper, we use the sum of the squared difference between the selected data and the solution of the model as the objective function to identify the transmission rate *β* and reporting rate *ρ* of each age group.

The data used are the daily reported cases from [3] during the dates of January 1 until February 28, 2021, dubbed the pre-vaccination period. For the objective function, we use the following three arbitrary differential equations

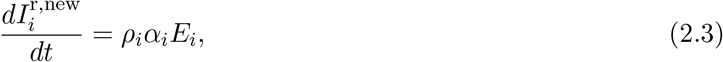

where 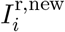 denotes the cumulative new reported cases for age group *i*. The differences of these solutions are the reported daily new cases. To incorporate the probable late reporting of cases, we consider the 7-day moving average as the base data. A derivative-free simplex method is used which is capable of constrained and multivariate optimization. Latin hypercube sampling [28, 35] is also done to better sample the initial guesses of the method.

Upon choosing the best fit among the estimated parameters, bootstrapping [18] is performed by redoing the parameter estimation multiple times with the best model fit as the data used including Poisson noise. By increasing the number of realizations, the estimated parameters may change and a confidence interval will be determined.

### 2.5 Optimal Control Problem

For the control problem in this study, we pay particular attention to vaccination efforts. Three controls are considered, each of which corresponds to the vaccination efforts for each age group. Specifically, the controls *u*_1_(*t*), *u*_2_(*t*), *u*_3_(*t*) denote the vaccination effort for the young, adult, and elderly age group, respectively. Incorporating these controls in Model (2.2) yields the following *disease-controlled model*

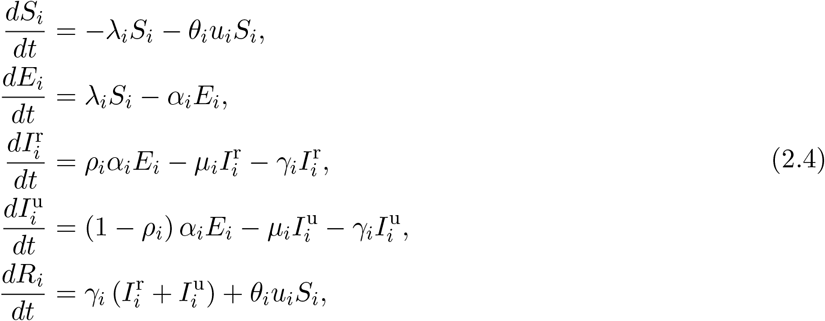

where *θ*_*i*_ denote vaccine efficacy for age group *i*. Note that an assumption of this system is that only susceptible individuals are candidates for vaccination. Here, the Pontryagin Maximum Principle (PMP) is employed to formulate the optimal control problem [34].

The objective is to minimize the number of infected individuals in 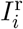 and 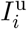 across different age groups with minimum vaccination administrative cost. This yields the following objective functional

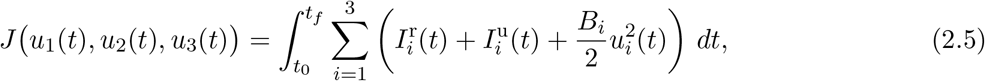

where [*t*_0_, *t*_*f*_] refers to the period of vaccination effort, and for *i* = 1, 2, 3, *B*_*i*_’s are weight constants representing the costs required to administer the corresponding control measures. Note that the controls are expressed as quadratic functions to take into account the nonlinear costs for the implementation of each control. Utilizing PMP leads to minimizing the objective functional in Equation (2.5) subject to the constraints given by Model (2.4).

We want to identify the optimal controls 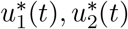, and 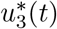 such that

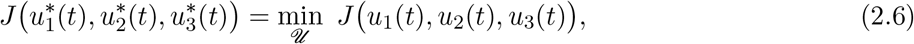

where

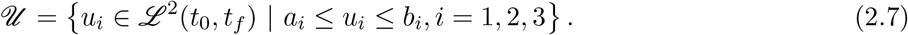

The upper (*b*_*i*_) and lower (*a*_*i*_) bounds for each control represent the maximum and minimum implementation efforts, respectively. If *u*_*i*_ = 0, then it suggests that no vaccination effort is employed, while *u*_*i*_ = 1 denotes that maximum effort is exerted for vaccine administration in the corresponding age group *i*. A theorem stating the existence of the optimal controls and corresponding states is given in Appendix B.

## 3 Results and Discussion

### 3.1 Estimated Transmission and Reporting Rates

To complete the parameters in Table 2.1, we use parameter estimation using new cases from January 1 up to February 28, 2021, to get the transmission and reporting rates for each of the three age groups. Figure 5 shows the best fit of the model compared to the data on cumulative cases for the young, adult, and elderly groups. From the parameter bootstrapping, the 95% confidence interval is also shown as bands around the best model fit. We close up as well on the cumulative data and model solution at the endpoint on February 28, 2021.

**Figure 5:**
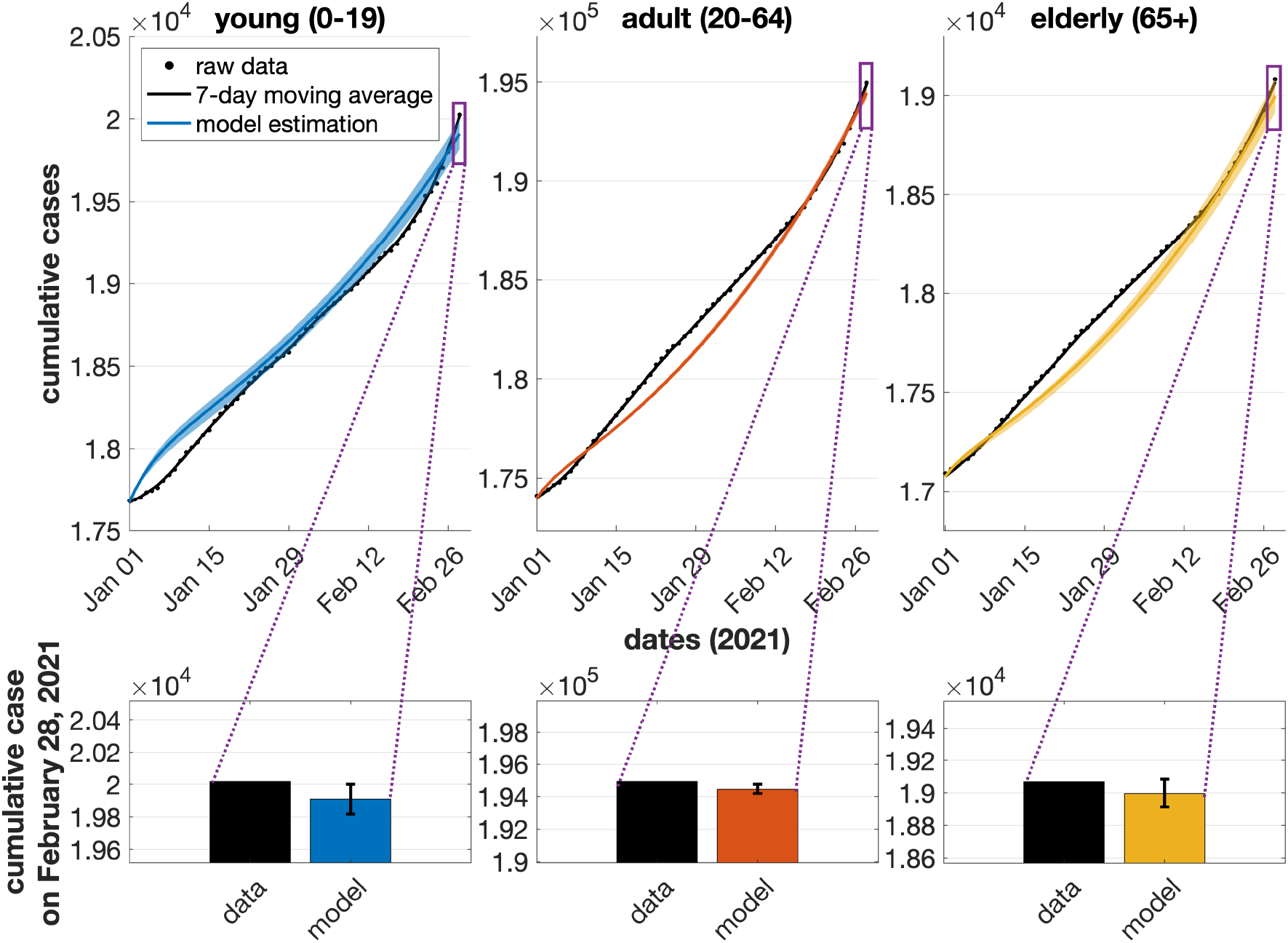
Comparison between raw data (black dots), 7-day moving average data (black line), and model output with 95% confidence interval for young (blue), adult (red), and elderly (yellow) compartments for cumulative data. Close-up (violet) of the cumulative data on the last day, February 28, 2021, is also shown for comparison between the data and the model.

Meanwhile, the estimated parameters and their mean and median values from the 1000 bootstrap realizations are presented in Table 3.1 and shown as distributions in Figure 6. Transmission is highest among the elderly with *β*_3_ = 0.0288 and least among the young age group with *β*_1_ = 0.0057. The mean reporting rate for the young, adult, and elderly are 22.36%, 70.14%, and 96.06%, respectively. Since the elderly are more likely to experience severe symptoms than the younger age groups, then we observe a higher reporting rate in the elderly group. Meanwhile, there is possible under-reporting in children and teenagers since testing resources are primarily given to priority groups such as front-line workers and symptomatic patients.

**Table 3.1:** Best parameter estimates for the transmission and reporting rates using the least-squares method, and the mean and median of the 1000 re-estimates in bootstrapping

| Parameter | Bootstrap Results |  |  |  |  |  |  |  |  |
| --- | --- | --- | --- | --- | --- | --- | --- | --- | --- |
|  | Young (0-19) |  |  | Adult (20-64) |  |  | Elderly (65+) |  |  |
|  | Initial | Mean | Median | Initial | Mean | Median | Initial | Mean | Median |
| transmission rate ($\beta$) | 0.0057 | 0.0057 | 0.0056 | 0.0115 | 0.0115 | 0.0115 | 0.0278 | 0.0288 | 0.0284 |
| reporting rate ($\rho$) | 0.2238 | 0.2236 | 0.2240 | 0.7006 | 0.7014 | 0.7011 | 1.000 | 0.9606 | 0.9980 |

**Figure 6:**
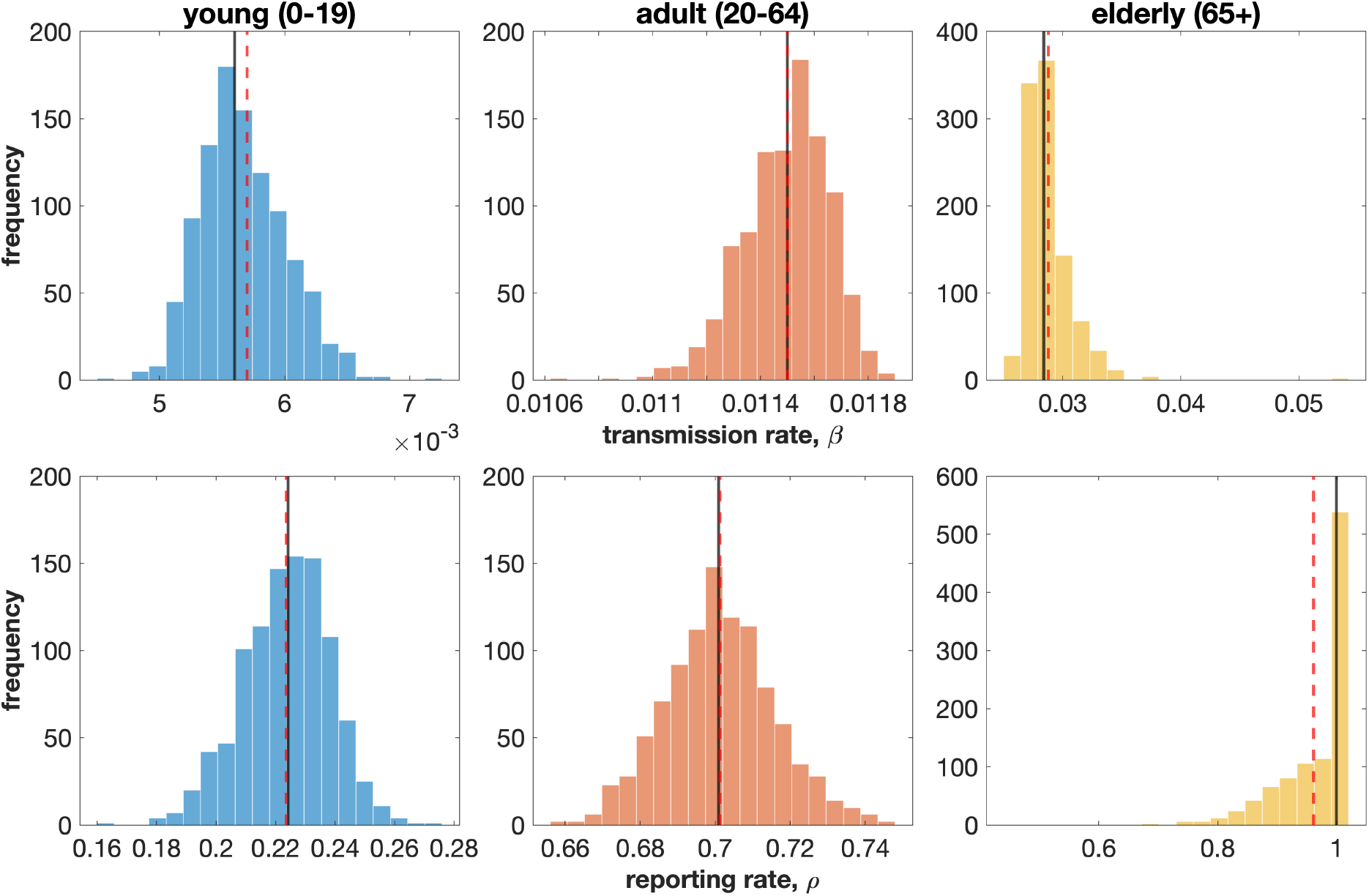
Histogram of parameter estimates for each age group after 1000 realizations from bootstrapping with their respective mean (red, dashed) and median (black, solid) values.

### 3.2 Optimal Vaccination Strategies

The main strategy used in this study is adding vaccination efforts according to the age-dependent vaccine prioritization imposed by the government. We use optimal controls for each age group to represent this pharmaceutical intervention. These optimal vaccination strategies are obtained by solving Model (2.4) with vaccine efficacy set to *θ*_*i*_ = 90% for all age groups, *i* = 1, 2, 3, and with weight parameters *B*_1_ = 10^*−*6^, *B*_2_ = 10^*−*6^, and *B*_3_ = 10^*−*5^, shown in Table 3.2.

**Table 3.2:** Values used for the numerical simulations of the optimal control problem for each time period and specific to each age group.

| Time Period | Age Group | Minimum daily vaccination rate | Maximum daily vaccination rate | Weight parameter |
| --- | --- | --- | --- | --- |
| Period 1<br>(March 1 - June 6, 2021) | 0-19 | 0 | 0 | $10^{-6}$ |
| | 20-64 | 0 | 0 | $10^{-6}$ |
| | 65+ | 0.001 | 0.01 | $10^{-5}$ |
| Period 2<br>(June 7 - October 14, 2021) | 0-19 | 0 | 0 | $10^{-6}$ |
| | 20-64 | 0.001 | 0.01 | $10^{-6}$ |
| | 65+ | 0.001 | 0.03 | $10^{-5}$ |
| Period 3<br>(October 15, 2021 - February 28, 2022) | 0-19 | 0.001 | 0.1 | $10^{-6}$ |
| | 20-64 | 0.001 | 0.1 | $10^{-6}$ |
| | 65+ | 0.001 | 0.1 | $10^{-5}$ |

A one-year period from March 1, 2021, the official starting date of vaccination in the Philippines, is chosen as the time window for the optimal vaccination strategies. We divide this vaccination period into three: first, March 1, 2021, to June 6, 2021, where the vaccine inoculation is only administered to the elderly [9]; second, June 7, 2021, to October 14, 2021, where the adult population, most of whom are in the working class, started to be inoculated [4]; and last, October 15, 2021, to February 28, 2022, where the young population was allowed to be vaccinated [7].

Furthermore, we assume that once vaccination becomes available to a particular age group, the maximum vaccination rate for the first period of implementation is low due to the adjustments in implementation, and speeds up in the next vaccination period. It is also assumed that in the third period, vaccine supply is sufficient so that all control parameters for vaccination can be increased to 0.1. Specifically, for the elderly population, the maximum vaccination rate in the first period is set at 0.01, increased to 0.03 in the second period, and increased further in the third period to 0.1. For the adult population, it is set at zero in the first period as no vaccination is available yet, then it is set at 0.01 in the second period, and then increased to 0.1 in the third period. Lastly, for the young population, the maximum vaccination rate is set at zero in the first two periods and is increased to 0.1 in the last period. Once vaccination becomes available to a certain age group, implementation is continuous even if they are not the priority group. Then, the lower bounds for the vaccination efforts are all set at a relatively lower value of 0.001. These additional values used for the optimal control problem are summarized in Table 3.2.

Figure 7 shows the optimal vaccination strategy obtained for all the age groups. The daily scheduled vaccination effort for the elderly (*u*_3_) shall be maintained at the upper bound for Periods 1 and 2. In particular, the optimal control chooses the maximum vaccination effort to prevent the elderly and the adult groups from getting infected. It suggests that efficient and effective implementation of vaccine administration, exhausting all possible limited resources. In Period 3, more vaccine supply is available, vaccine administration is easier, and vaccination hesitancy is lower than the previous periods. However, the results depict that the vaccination efforts for all the groups should be at the maximum for the first few months and must be sustained for the adult and young groups throughout the considered inoculation duration. It is more likely that the target number of elderly to be inoculated is achieved around the midway of Period 3. Thus, the control effort for the elderly can be decreased and then re-allocated to the other groups.

**Figure 7:**
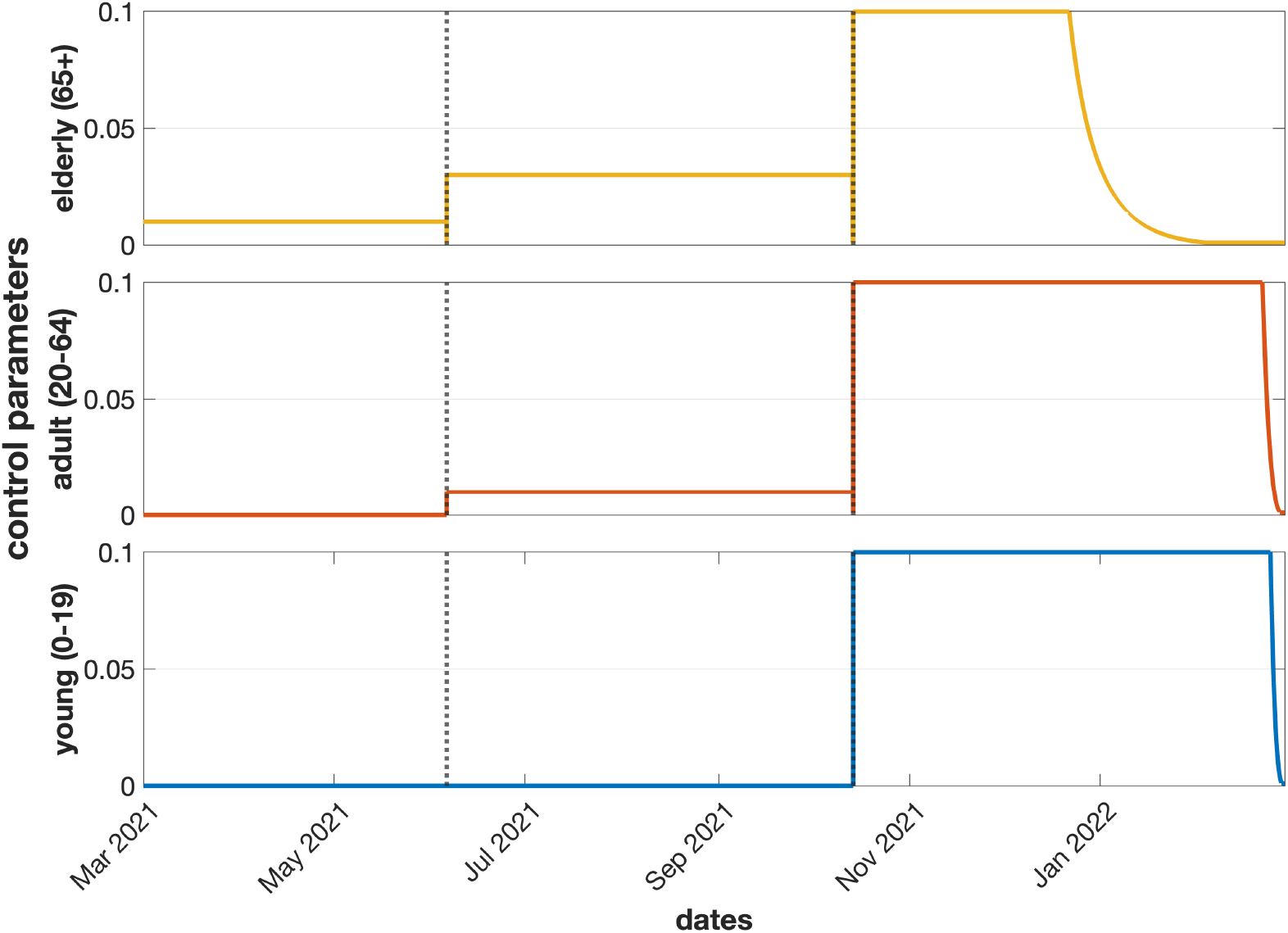
Control profiles for the vaccination of the elderly, adult, and young populations for Period 1 (March 1, 2021 - June 6, 2021), Period 2 (June 7, 2021 - October 14, 2021), and Period 3 (October 15, 2021 - February 28, 2022).

For the daily scheduled vaccination effort for the adults (*u*_2_), once they are allowed to be vaccinated (from June onward), a maximum daily vaccination effort shall be scheduled in Period 2. The maximum effort shall remain for Period 3 until before the end of February when vaccination can be lowered.

Finally, the daily scheduled vaccination effort for the younger population (*u*_1_) shall also be at maximum, once they are allowed to be vaccinated (from October 15 onward). Vaccination can be decreased towards the end of February, a little later than the lowering of efforts for the adult population. Generally, throughout the course of the first two periods, in order to reach optimal results, the vaccination efforts must be set to the maximum value. For the third period, the control efforts can be lowered towards the end with a relatively earlier lowering of vaccination efforts for the elderly population which can be attributed to their comparatively smaller population.

Figure 8 shows that without intervention, the projected number of daily infected cases can reach up to 21, 208 for the elderly population, 317, 640 for the adult population, and 128, 509 for the young population. Meanwhile, the optimal vaccination strategies result in a significant decrease in the number of infected individuals across all age groups. The controls result in a peak of 63, 753 cases for the adult age group, 28, 177 for the young age group, and 1, 537 for the elderly age group. It can also be observed that even without vaccination in the period June to October, the age group 0 *−* 19 exhibits a significant decrease in the infected population. This means that the vaccination of the adult and elderly populations have a great impact on reducing the number of infections in the younger population. It is observed from Figure 8 (left panel) that the number of daily incidence cases does not decrease immediately within the initial phase of vaccination roll-out, e.g., first and second period for elderly and adult, respectively. This can be attributed to the delayed immunization effect of the vaccines. Though the young individuals are vaccinated in the third period, the incidence cases are already decreasing due to the vaccination of elderly and adult population in the previous periods.

**Figure 8:**
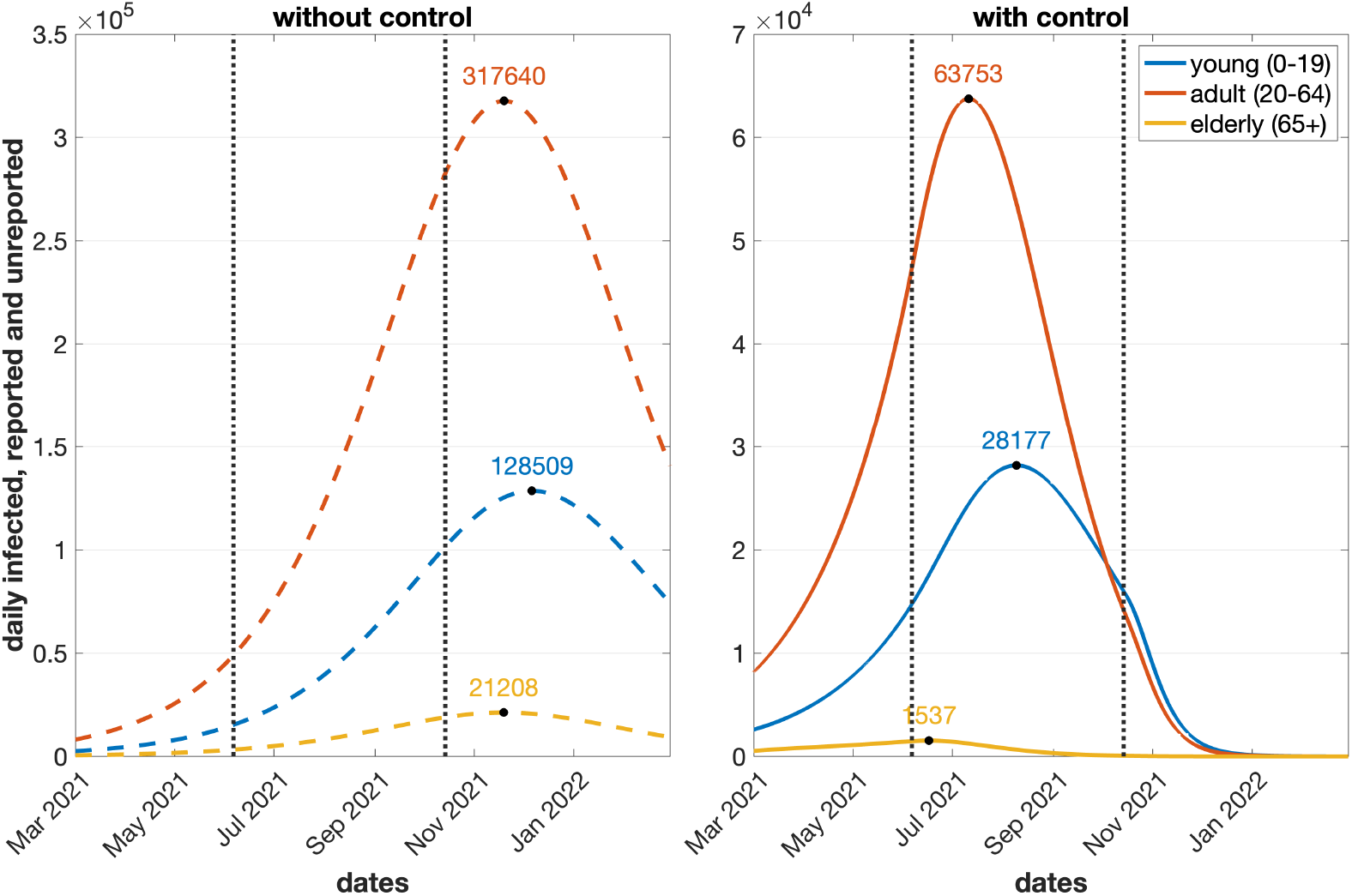
Total daily active cases without (dashed) and with (solid) optimal control strategies for both reported and unreported compartments for young, adult, and elderly populations.

Figure 9 shows a comparison of the infected populations with and without control. Even with control, the largest contributor to the total infections is the adult age group which is followed by the young population and then the elderly population. This is intuitive since the adult population comprises the bulk of the total population, and also the most mobile and interacting group. The inoculation of the elderly population during the first period led to a decrease in cumulative cases for all age groups, ranging from 41% for the young, 43% for the adult, and 49% for the elderly. When adults were allowed to get vaccines during the second period, a massive decrease in infections can be seen from uncontrolled to the controlled system. The percent decreases are 397% for the elderly, 251% for the adult, and 254% for the young. However, inoculation for the young population in the third vaccination period decreased this even further wherein a 392% drop can be observed for the young, 440% for the adult, and 897% for the elderly.

**Figure 9:**
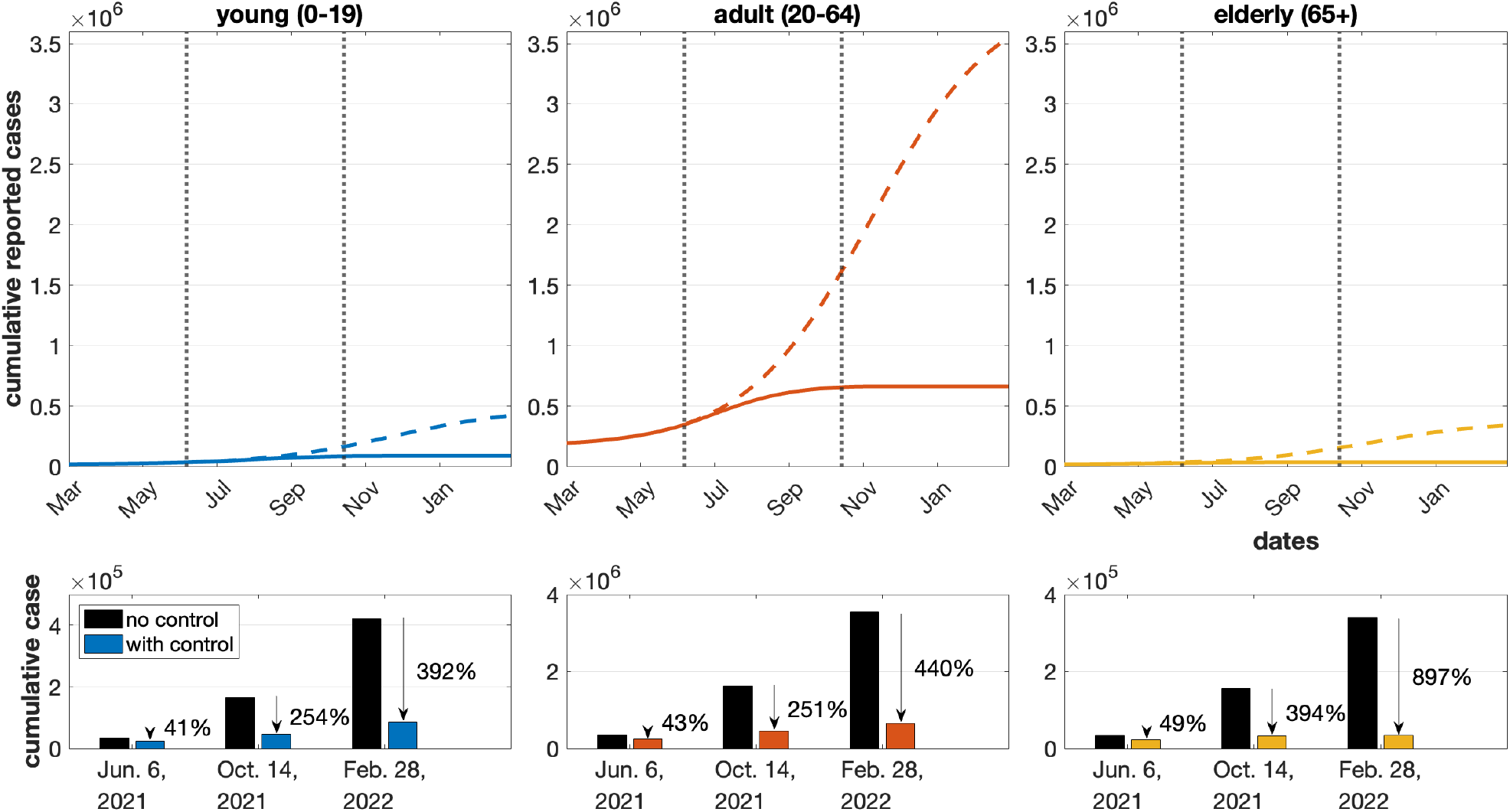
Upper panel shows the cumulative new reported COVID-19 cases without control (left panel, dashed) and with control (right panel, solid) for the young (blue), adult (red), and elderly (yellow). Lower panel depicts the cumulative cases at the end of each vaccination period and their respective percent decreases.

## 4 Conclusion

In this paper, we developed an age-structured compartmental model for COVID-19 with an unreported infectious class. The epidemiological compartments are divided into three groups: *young* (0-19 years), *adult* (20-64 years), and *elderly* (65+ years), according to vaccine prioritization. We denote these groups by *i* = 1, 2, and 3, respectively.

A contact matrix is used to incorporate the interaction among individuals across various age groups. Specifically, each entry of the matrix takes into account the contributions from the number of daily contacts between the considered age group *i* with other age groups *j* in household, workplace, school, and community. Furthermore, we introduced the force of infection denoted by *λ*_*i*_ for a susceptible individual for the *i*th age group. It is composed of two factors. The first one is the transmission rate for this particular age group. The second one is the sum of the product of the associated entry of the contact matrix and the proportion of the infected cases over all three age groups *j* = 1, 2, 3, for the given specific age group *i*. Moreover, the unreported cases are multiplied by a factor denoted by *σ*_*i*_, which is the relative contribution to transmission of the unreported class for the associated age group.

Most of the model parameters are obtained from the literature or aggregated from available data. Reporting and transmission rates are estimated because they are country/region-specific and are not readily available. Estimation was performed using a least-squares formulation by fitting the model to the cumulative case data. Results show that both reporting and transmission rates are higher in the older age group. Our simulation suggests that thousands of cases in young and adult populations were undetected. Uncertainty analysis showed the reliability of the estimates.

Given the limited availability and administration of the vaccines in the first half of the year 2021, we analyzed the effect of age-targeted vaccine distribution by comparing the model output when no intervention is imposed, against the output when vaccination is introduced in the model. The results of the numerical simulations for the controls show that efforts from the government in implementing its vaccination drive in the country should always be at maximum until towards the end of the last period when vaccination can be lowered initially for the elderly population first, then for the adult population, and lastly, for the young population. Moreover, implementing the vaccine strategies as early as possible can reduce the cumulative reported cases in the young, adult, and elderly population by 392%, 440%, and 897%, respectively. The results from the optimal control problem can be attributed to the proportion of each age group to the total population, the transmission rate specific to each age group, and the order of prioritization in vaccination.

Our model took into account age structure and unreporting. The model was applied when the vaccination program started, assuming that one vaccine dose can already provide immunity. Further model iterations are needed to include the impact of the succeeding shots and the effect of waning immunity. At the beginning of the vaccination period, only one variant was dominant in the Philippines so variants were also not incorporated into the model. Nevertheless, integrating these into the model is an exciting research direction, but would require other vaccination and breakthrough infection data and would demand a separate study. Ultimately, the aim of this study is to guide the policymakers in crafting strategies that can mitigate the spread of an infectious disease considering age-dependent transmission and limited vaccine supply. Although the model is tested using Philippine data, the model is general enough that it can be applied to other regions with similar situations.

## Data Availability

All data produced in the present study are available upon request to the authors.

## Acknowledgement

This project was supported by a grant from the University of the Philippines Computational Research Laboratory.

## Appendix A. Computation of the 3 *×* 3 Contact Matrices

The four 16 *×* 16 pre-pandemic contact matrices for different interactions and their total can be reduced to a size that clumps together the five-year age levels into three main age groups for the young, adult, and elderly populations. Let *C* be the corresponding 3 *×*3 total contact matrix from *D*. The superscripts also follow for reduced contact matrices for different interactions, e.g., *C*^*h*^ is the reduced *D*^*h*^.

The young age group contains ages from 0 to 19 that correspond to the first four age levels. The adult age group covers the most age levels which are from 20 to 64. Lastly, ages 65 and above will be classified under the elderly age group. From here, we can determine sets containing the indices of each of these age groups, i.e.,

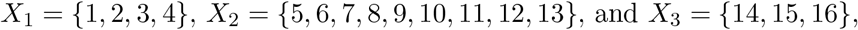

where *X*_1_ is for the young age group, *X*_2_ is for the adult age group, and *X*_3_ is for the elderly age group. This can be observed on Table .1.

**Table 1:** Age groups and their respective population fractions based on the 2015 NCR Population.

| Age group | Index ($i$) | Ages | Population per age group | Population fraction ($d_i$) |
| --- | --- | --- | --- | --- |
| Young | 1 | 0-4 | 1200443 | 0.093221978 |
|  | 2 | 5-9 | 1163719 | 0.090370128 |
|  | 3 | 10-14 | 1135592 | 0.088185889 |
|  | 4 | 15-19 | 1253803 | 0.097365719 |
| Adult | 5 | 20-24 | 1349005 | 0.104758756 |
|  | 6 | 25-29 | 1265102 | 0.098243158 |
|  | 7 | 30-34 | 1104618 | 0.085780562 |
|  | 8 | 35-39 | 963867 | 0.074850358 |
|  | 9 | 40-44 | 817789 | 0.063506479 |
|  | 10 | 45-49 | 712253 | 0.055310942 |
|  | 11 | 50-54 | 591442 | 0.045929206 |
|  | 12 | 55-59 | 468406 | 0.036374683 |
|  | 13 | 60-64 | 345164 | 0.026804164 |
| Elderly | 14 | 65-69 | 223238 | 0.017335840 |
|  | 15 | 70-74 | 122964 | 0.009548931 |
|  | 16 | 75+ | 159848 | 0.012413206 |
| Total |  |  | 12877253 | 1 |

Using the latest available data for the National Capital Region from the Philippine Statistics Authority [8], we can create a 16 *×* 1 vector *d* = (*d*_*k*_) that contains the population fraction *d*_*k*_ of each of the age level *k* for *D*. We determine its elements by dividing the age group population over the total population. The values of population fraction *d*_*k*_ for *k* = 1, 2, …, 16 are shown in Table .1.

The formula of obtaining *C* = (*C*_*ij*_) is

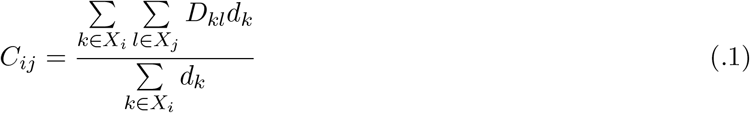

where *i, j ∈ {*1, 2, 3*}* correspond to the young, adult, and elderly populations, respectively.

For example, if we want to get *C*_13_ from the total contact matrix *D*, which is the contact value of the young population relative to the elderly population, we consider the indices *i ∈ X*_1_ and *j ∈ X*_3_. The denominator simply adds up the population fraction of the young population to get 0.3959. Meanwhile, the numerator considers the values for the 4 *×* 3 submatrix of *D* with row indices from *X*_1_ and column indices from *X*_3_. Each row is summed and multiplied by its respective population fraction. For this example, we will get the numerator as 0.2110. Hence, the value of *C*_13_ is 0.533. This can be observed as the 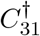 of the first matrix plot in Figure 10.

**Figure 10:**
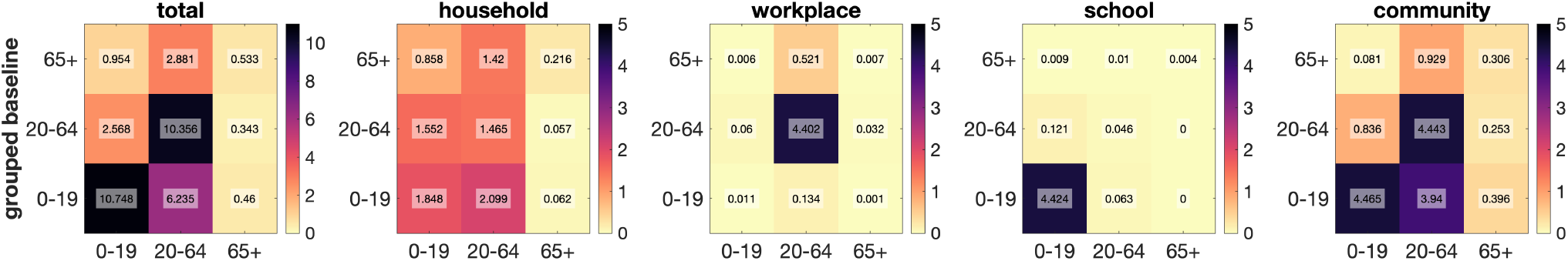
The obtained 3× 3 contact matrices considering the young, adult, and elderly population groups in total, household, workplace, school, and community context. For visualization, the shown matrices *C*^*†*^ are flipped upside down, i.e., *C*^*†*^ = 𝕁*C* where𝕁is the backward identity.

## Appendix B. Existence of Optimal Control and Adjoint Equations

### Theorem .1.

*There exist optimal control* 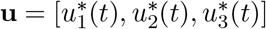 *and corresponding optimal state* 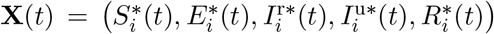 *for i* = 1, 2, 3, *which minimize the objective functional* (2.5) *over all controls in* (2.7). *Given this optimal pair of solution* (**u, X**), *there exist adjoint functions ψ*_*k*_, *k* = 1, 2, …, 15 *such that for the Hamiltonian given by*

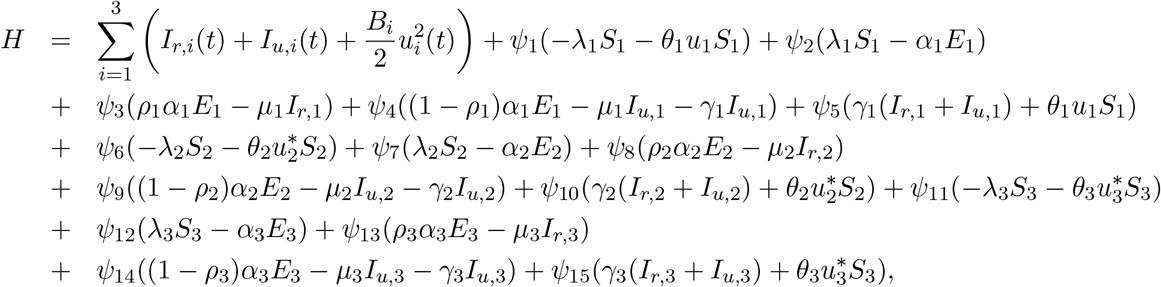

*we have*

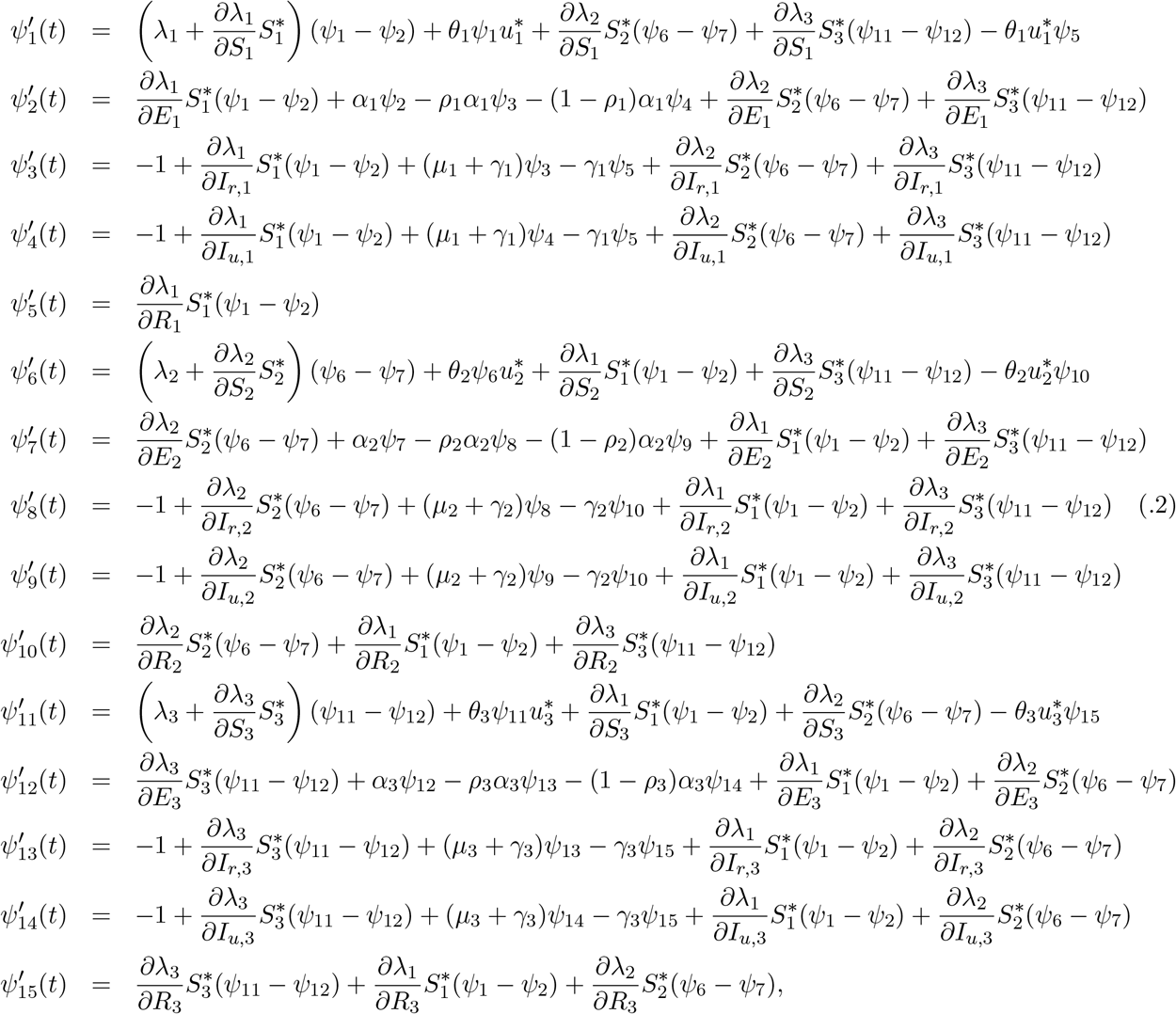

*with transversality conditions*

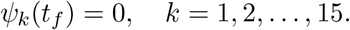

*Furthermore*,

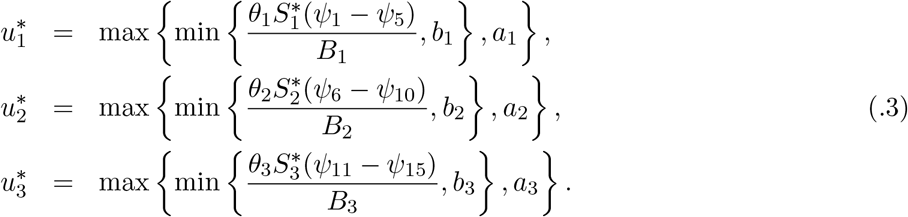

*Proof*. The optimal control 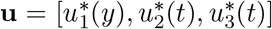 satisfying Equation (2.6) exists due to the convexity of the integrand in Equation (2.5). We use Pontryagin’s Maximum Principle [38] to obtain the adjoint equations and transversality conditions. Differentiating the Hamiltonian *H* with respect to the state variables gives us the following system of equations:

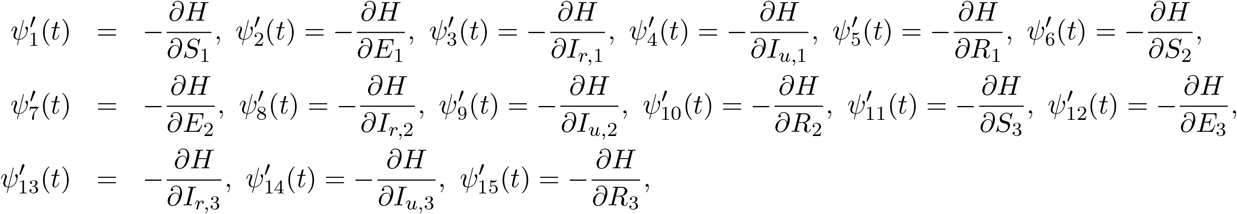

resulting to System (.2) with *ψ*_*k*_(*t*_*f*_) = 0 for *k* = 1, 2, …, 15.

Moreover, we derive 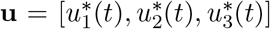 on *U* by solving the optimality conditions 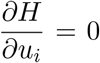 for *i* = 1, 2, 3, i.e.,

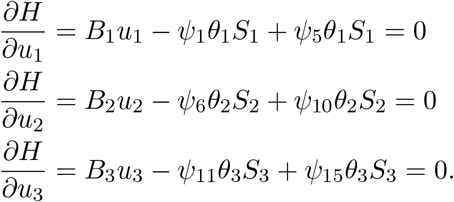

Evaluating at 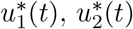, and 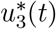 on 𝒰, we get

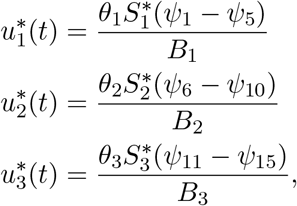

and by considering the upper (*b*_*i*_) and lower (*a*_*i*_) bounds for each control, we end up with Equations (.3).

